# Quantitative analysis of insecticides in long-lasting insecticidal nets using liquid chromatography mass spectrometry (LC-MS) and X-ray fluorescence (XRF) spectroscopy

**DOI:** 10.1101/2023.03.06.23286872

**Authors:** Melanie Koinari, Nakei Bubun, David Wilson, Evodia Anetul, Lincoln Timinao, Petrina Johnson, Norelle Daly, Moses Laman, Tim Freeman, Stephan Karl

## Abstract

**Background:** Long lasting insecticidal nets (LLIN) are a key vector control tool used for the prevention of malaria. Active ingredient (AI) measurements in LLIN are essential for evaluating their quality and effectiveness. The main aim of the present study was to determine the utility of X-ray fluorescence (XRF) spectroscopy as a suitable in-field tool for total AI quantification in LLINs.

**Methods:** New and unused LLIN samples containing deltamethrin (PermaNet® 2.0, n = 35) and alpha-cypermethrin (SafeNet®, n = 43) were obtained from batches delivered to PNG for mass distribution. Insecticides were extracted from the LLINs using a simple extraction technique and quantified using liquid chromatography mass spectrometry (LC-MS). The LC-MS results were correlated with in-field XRF spectroscopy measurements on the same nets. Operators were blinded towards the identity of the nets. Bioefficacy of the LLIN samples was tested using WHO cone bioassays and test results were correlated with total AI content.

**Results:** The results indicate a close agreement between the quantitative XRF and LC-MS. Interestingly, the total AI content was negatively correlated with bioefficacy in PermaNet® 2.0 (especially, in recently manufactured nets). In contrast, AI content was positively correlated with bioefficacy in SafeNet®. These results indicate that the chemical content analysis in predelivery inspections does not always predict bioefficacy well.

**Conclusion:** XRF is a promising in-field method for quantification of both deltamethrin and alpha-cypermethrin coated LLINs. Since total AI content is not always a predictor of the efficacy of LLINs to kill mosquitoes, bioefficacy measurements should be included in predelivery inspections.

## Background

Hundreds of millions of long lasting insecticidal nets (LLINs) for vector control are procured annually by international donors and distributed [1]. LLIN distributions have helped to reduce the malaria burden in many endemic countries [2]. Despite this, and the target to eliminate malaria globally, malaria case numbers have stagnated in the last decade [3]. Mass distribution of LLINs is the backbone of malaria vector control in Papua New Guinea (PNG). About 14 million bed nets have been distributed in PNG since 2009 [4]. Bed nets are distributed every 3 years to all villages in PNG except above 1600 meters, using a ‘rolling’ distribution schedule to maintain sustainability. Previous studies in PNG found that bed net performance significantly decreased in 2013 following a manufacturing change involving the chemical coating of the nets leading to reduced efficacy of the nets to kill mosquitoes and thus reduced community protection [5-7].

Many previous studies have shown that bioefficacy in used LLINs is strongly correlated with total insecticide content [8-10]. This is normal because when nets are washed or exposed to UV radiation (sun exposure), total active ingredient (AI) content decreases and once a threshold AI concentration is reached, 100% bioefficacy normally observed with fully susceptible strains in new and unwashed nets is not maintained. However, other studies have demonstrated that recently manufactured new and unused LLIN products may not be able to pass WHO cone bioassay performance criteria with susceptible mosquito colonies, even though their total AI content was determined to be adequate in predelivery inspections [7, 11]. This may be for different reasons including restricted bioavailability of AI on the net surface due to the nature of the LLIN coating, presence of AI in chemical or physical states that may be detrimental to its effectiveness (such as isomers with reduced potency or crystalline states that may reduce the uptake of AI by mosquitoes) [5, 6, 12, 13]. Thus, it is important to fully understand the relationship between LLIN bioefficacy as determined in simple and standardized evaluations e.g., using WHO cone bioassays, and total AI concentration and presentation on the surface of LLINs.

Prior to 2009, only three LLINs were recommended by World Health Organization (WHO): a deltamethrin-coated net, a permethrin-incorporated net and an alpha-cypermethrin-coated net [14]. As of 2017, there are 23 prequalified LLINs, manufactured by 14 companies [15]. Spread of pyrethroid resistance necessitated the development new products with alternative (active ingredients (AI) formulations, leading to further diversification of the available product palette [16].

Chemical analyses of the LLIN forms an integral part of the WHO guideline for testing new nets as well as monitoring the durability of LNs under operational conditions [17]. As stated in the guideline, the insecticide content of a net samples should be analyzed by the methods published by the Collaborative International Pesticides Analytical Council (CIPAC), specifically, the high-pressure liquid chromatography (HPLC) [18-20]. However, laboratories in malaria endemic developing countries that evaluate LLINs for malaria control and prevention or conduct quality assurance tests for national programs may not have easy access to the equipment needed or the expertise required. As such, simple, in-field approaches to quantify insecticides in LLIN products would be useful. A number of alternative methods have been developed to quantify pyrethroids in LLINs including rapid colorimetric field tests and various quantitative and imaging techniques [21–23].

One of the most promising alternative methods is x-ray fluorescence (XRF) spectroscopy. Researchers in Ghana, Ethiopia and Guatemala have demonstrated the utility of XRF for fast quantification of deltamethrin in the field, specifically, for durability assessment of PermaNet® LLINs after (3 -38) months of use [8, 24, 25]. While XRF has been shown to be able to quantify deltamathrin, little is known if this concept has been applied to other insecticides even though this is theoretically possible using the same methodology.

The present study served several main purposes. Firstly, to correlate total insecticide content measured using a laboratory-based method (liquid chromatography mass spectrometry) and a field-based method (XRF spectroscopy) for both deltamethrin and alpha-cypermethrin to determine the utility of XRF as a suitable in-field tool for total AI quantification for both of these insecticides, which are found in the vast majority of prequalified LLIN products. Secondly, to determine the correlation between total AI content and bioefficacy in two LLIN products (one deltametrin product and one alpha-cypermethrin product) delivered to PNG for mass distribution in the new and unused state.

### Methods

WHO cone bioassays and XRF spectroscopy were conducted at the Vector-borne Diseases Unit of the Papua New Guinea Institute of Medical Research (PNG IMR) while the LC-MS was conducted at the Australian Institute of Tropical Health and Medicine (AITHM), James Cook University (JCU), Cairns.

### LLIN sampling

PermaNet® 2.0 (n = 35) and SafeNet® (n = 45), all unused and in their original packaging, were included in the present study. As described in Vinit et al., 2020, unused PermaNet® 2.0 manufactured in 2007 to 2017 were obtained from villages or provincial health authorities in various provinces in PNG. Rotarians Against Malaria (RAM) PNG provided PermaNets® 2.0 manufactured in 2018 and 2019 from consignments dedicated to different provinces [7]. SafeNet® LLINs manufactured in 2019 and 2020 were provided by RAM PNG. Details of the selected nets can be found in Additional file 1: Table S1. PermaNet 2.0 samples and SafeNet samples used in the LCMS, XRF and cone bioassay analysis.

### X-ray fluorescence analysis on single layers of netting

Proxy element (bromine and chlorine) content was measured on a monolayer of netting, using a Vanta® Handheld XRF Analyser (Olympus, Australia). To increase the limit of detection the instrument was used with a shielded measurement chamber and silica standard background, also sourced from Olympus. Bromine and chlorine were quantified by the built-in software and expressed in parts-per-million (ppm). A measurement took about 45 seconds.

## Liquid Chromatography Mass Spectrophotometry (LC-MS)

### Insecticide extraction

C-MS analysis was performed on methanol extracts of two samples of each net (each piece 2 cm x 2 cm), which were extracted separately in duplicate to generate technical and biological replicates. Net samples were weighed and submerged in 1 mL of LC-MS-grade methanol (Merck, Australia) in individual 2.0 mL Eppendorf Safelock tubes (Eppendorf, Australia) previously shown to be free of any contaminants (e.g., plasticizer from manufacture). The tube was placed on a vortex for 60 s then briefly centrifuged.

### LC-MS measurement

LC-MS was performed using a Shimadzu LC-MS2020 (Shimadzu, Japan) and a Phenomenex Lux cellulose-2 chiral column, (150 × 2.0 mm; Phenomenex, Australia) at 40°C, and 5 mM ammonium acetate/water (Solvent A) and 100 % methanol (Solvent B; LiChrosolv LC-MS-grade, Merck) mobile phase at 0.25 mL/min flow rate. Samples (20 μL) were eluted isocratically in 82% Solvent B over 12 min and UV absorbance monitored at 246 and 280 nm. Mass spectra were collected in positive ion mode over a scan range of m/z 200-800 with a detector voltage of 1.15 kV, nebulizing gas flow of 1.5 L/min, and drying gas flow of 3.0 L/min.

### Standards

Analytical grade samples of deltamethrin and alpha-cypermethrin, to be used as standards, were purchased from Merck (Australia). Standard curves that spanned the anticipated insecticide concentration range for each LLIN product (for PermaNet® 2.0, target deltamethrin is 1.8 g/kg and for SafeNet®, 5g/kg alpha-cypermethrin) were created by preparing dilution series of known insecticide concentrations in methanol. A primary stock solution was prepared by dissolving an accurately weighed amount of insecticide on an analytical balance (Mettler Toledo, Australia) in 1 mL methanol. The stock was then diluted to 5x the expected target concentration, followed by a 1.5 fold dilution series for a total of ten dilution samples of the standard insecticide. These known concentrations of standard insecticides were included in every LC-MS analysis of net samples (with unknown insecticide content).

### Extraction Protocol Validation

In order to determine efficient extraction of the insecticides from the nets, a subset of net samples was subjected to the following validation experiments. First, in solvent volume experiments: n = 7 samples cut from each LLIN (2012-PermaNet 2.0, n=2) were submerged in 500, 750, 1000, 1200, 1500, 1750 and 2000 μL of methanol. Extracts from each volume were analysed by LC-MS in triplicates. Second, extraction time; n = 5 samples cut from each LLIN (2012-PermaNet 2.0, n=2) were each extracted in 1000 μL of methanol. The 5 samples were placed in the solvent at the same time but were removed at 10 mins (0 h), 1 h, 3 h, 5 h and 24 h. Extracts from each extraction time point were analysed by LC-MS in triplicate. Third, in order to validate complete extraction from the net samples, n = 4 samples were cut from an LLIN (2020 SafeNet®, n = 1), were repeatedly (twice) subjected to the above extraction process (solvent volume = 1000 μL; extraction time = 10 mins, vortex = 1 min and a brief centrifuge).

### WHO Cone Bioassays

Cone bioassays were performed following the WHO guidelines and under laboratory conditions with 28.5 ± 2.7 °C and 65 ± 9 % relative humidity [17]. Using an aspirator, 5 insecticide susceptible, non-blood-fed, 2-5 day old female *Anopheles farauti* mosquitoes were introduced into a cone and a cotton ball was used to plug the hole. Mosquitoes were exposed to the net pieces for 3 min (timed individually for each cone), after which they were gently transferred from the cones to a holding cup screened with untreated netting and provided access to 10% sugar solution via a soaked piece of cotton wool placed on top of the netting. Knockdown was recorded at 60 min after exposure and mortality was recorded at 24 h after exposure.

### Data analysis

Absorbance and chromatogram data from LC-MS are exported to Microsoft Excel 2016 (Microsoft Inc.) and GraphPad Prism 9.3.1 (GraphPad Software) for analyses. The absorbance peaks for the known concentrations in the insecticide standards were used to plot the standard curve. The standard curve was then used to determine the insecticide concentration in the unknown samples in g/kg.

## Results

### LC-MS protocol validation experiments

Solvent volume and extraction time did not affect the experimental outcomes systematically. For all nets used in these validation experiments, the measured deltamethrin concentrations were in the expected range and between 1.7 - 1.9 g/kg (Figure 1A and B). In all further analyses, 1 mL of methanol was used and extraction time was fixed to 10 mins. Additionally, repeated extractions indicated that the efficiency of the first extraction was >95% (Figure 1C).

**Figure 1.**
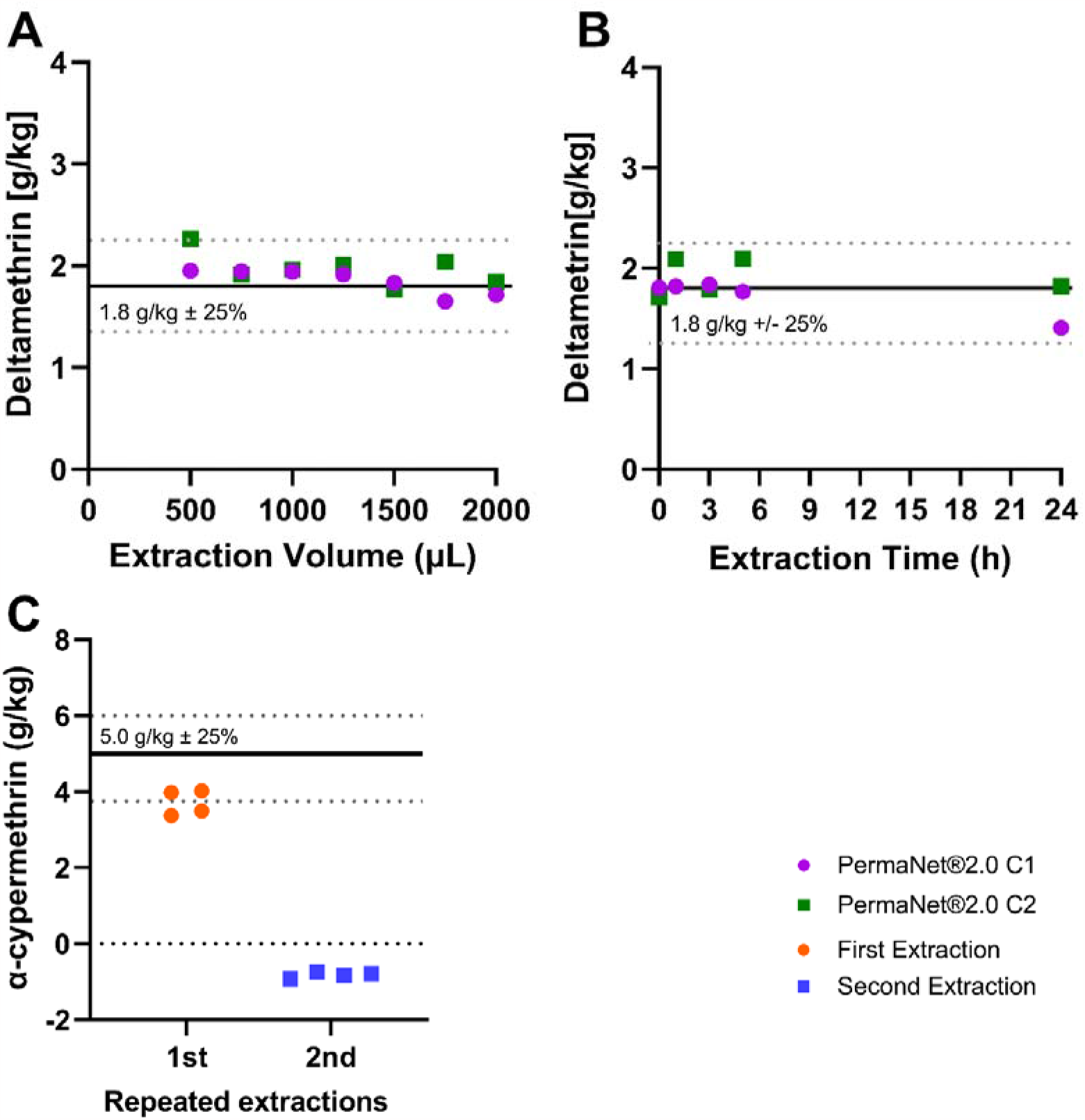
Extraction Efficiency. **Panel A** shows the concentration 190 of deltamethrin for n=7samples (from each 2012-PermaNet®) extracted at various volume of methonol. **Panel B** shows the concentration of deltamentrin for the n=5 samples (from each 2012-PermaNet®) extracted at different time points. **Panel C** shows the concentration of α-cypermethrin for the n = 4 samples (from a 2020-SafeNet®) which were extracted at 10 mins in 1 mL of methanol. Horizontal lines indicate the target AI concentrations as per product label (average +/-25%).

### Correlation of insecticide content measurements between XRF and LC-MS methods

XRF and LC-MS insecticide quantification results were compared for both PermaNet® 2.0 and SafeNet® LLINs (Figure 2A and B). XRF and LCMS data were highly significantly correlated for both products (PermaNet® 2.0 R=0.77, p<0.0001 and Safenet ® R=0.23, p=0.0012, respectively), however the correlation was much better for Permanet 2.0. Batches of products clustered significantly indicating differences between specific product batches. In particular, for PermaNet® 2.0 this may be due to the wide range of manufacture years dating back to 2007. PermaNet® 2.0, older nets generally contained less AI (p<0.0001), indicating that AI may have been gradually lost over time. Bland and Altman analysis (presented in Figure 2C and D) indicated good agreement between XRF and LCMS quantification for both products, with no systematic bias. The 95 % confidence of agreement between methods were substantial.

**Figure 2:**
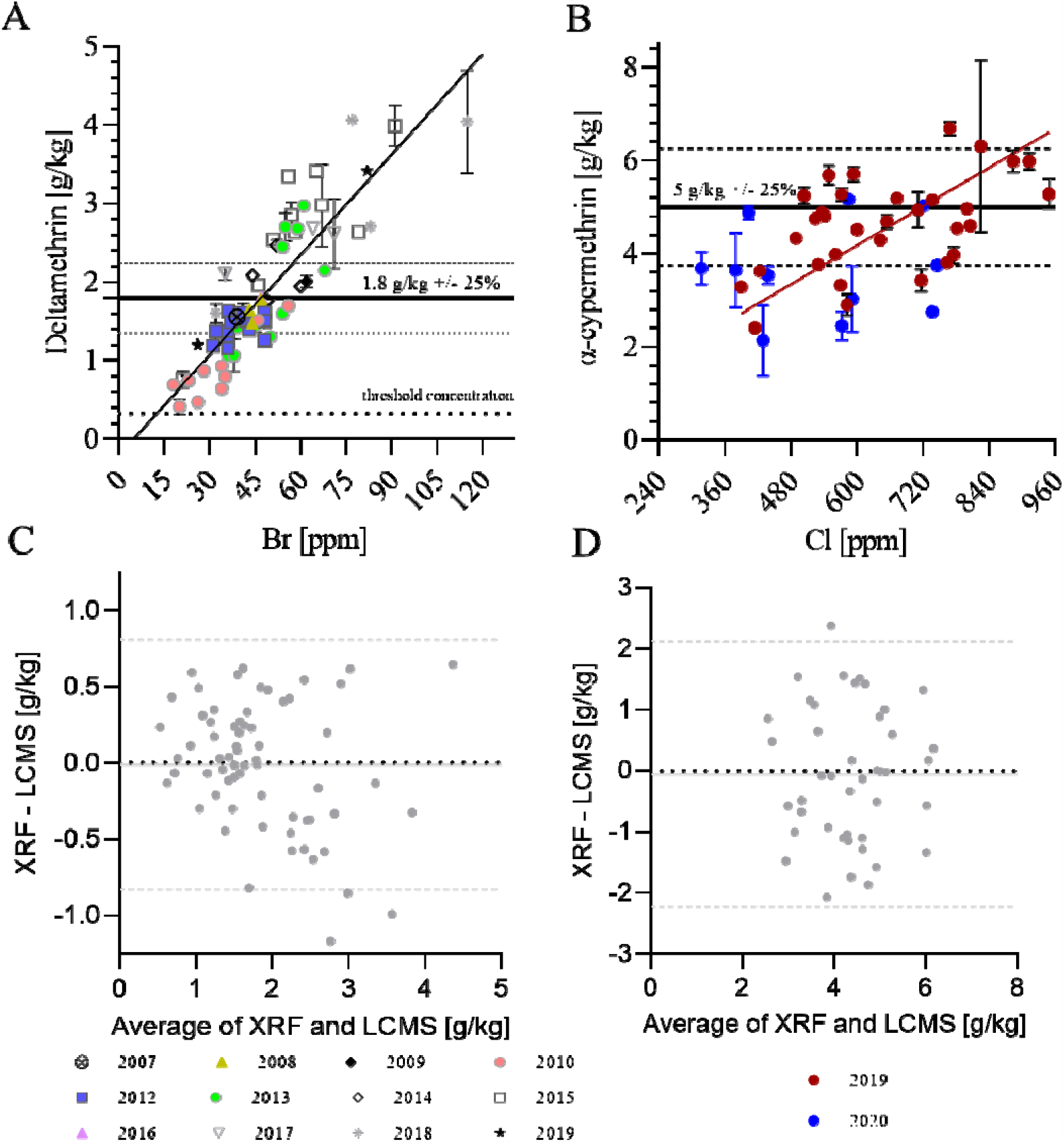
Correlation of total AI quantification using XRF and LC-MS techniques. **Panel A:** Correlation of XRF (in ppm) and LC-MS (in g/kg) data for PermaNet® 2.0 samples with manufacturing years between 2007 and 2019. **Panel B:** Correlation of XRF and LC-MS data for SafeNet® samples with manufacturing years between 2019 and 2020. Horizontal lines in Panels A and B indicate the target concentrations for the products as per product label. Simple linear regression curves are also shown. The regression was used to convert the XRF data in ppm to the corresponding g/kg. **Panels C and D:** Bland and Altman analyses for XRF versus LC-MS quantification of total AI content (Panel C - Permanet 2.0 ®; Panel D - Safenet ®). The plots show the differences XRF/LC-MS data pairs over their averages. Horizontal lines in Panels C and D indicate average bias (continuous line) and 95% confidence levels of agreement (dashed lines).

### Correlation of Insecticide Content with Bioefficacy

The correlation between the results of standardised WHO cone bioassays and the chemical analysis results are shown in Figure 3. Strikingly, bioefficacy was lower for more recent (2013-2019) PermaNet® 2.0 samples with higher total AI content (R=-0.57, p<0.0001). This observation illustrates clearly, that total AI content is not a robust indicator for bioefficacy. In contrast, a positive correlation was observed between bioefficacy and total AI content in the SafeNet® samples. This is normally expected in particular with used nets, but in this case still surprising as the samples in the present study were from new and unused nets, and it was expect that they would all exhibit 100% mortality as indicated in the respective WHOPES report underlying prequalification of SafeNet® [26].

**Figure 3:**
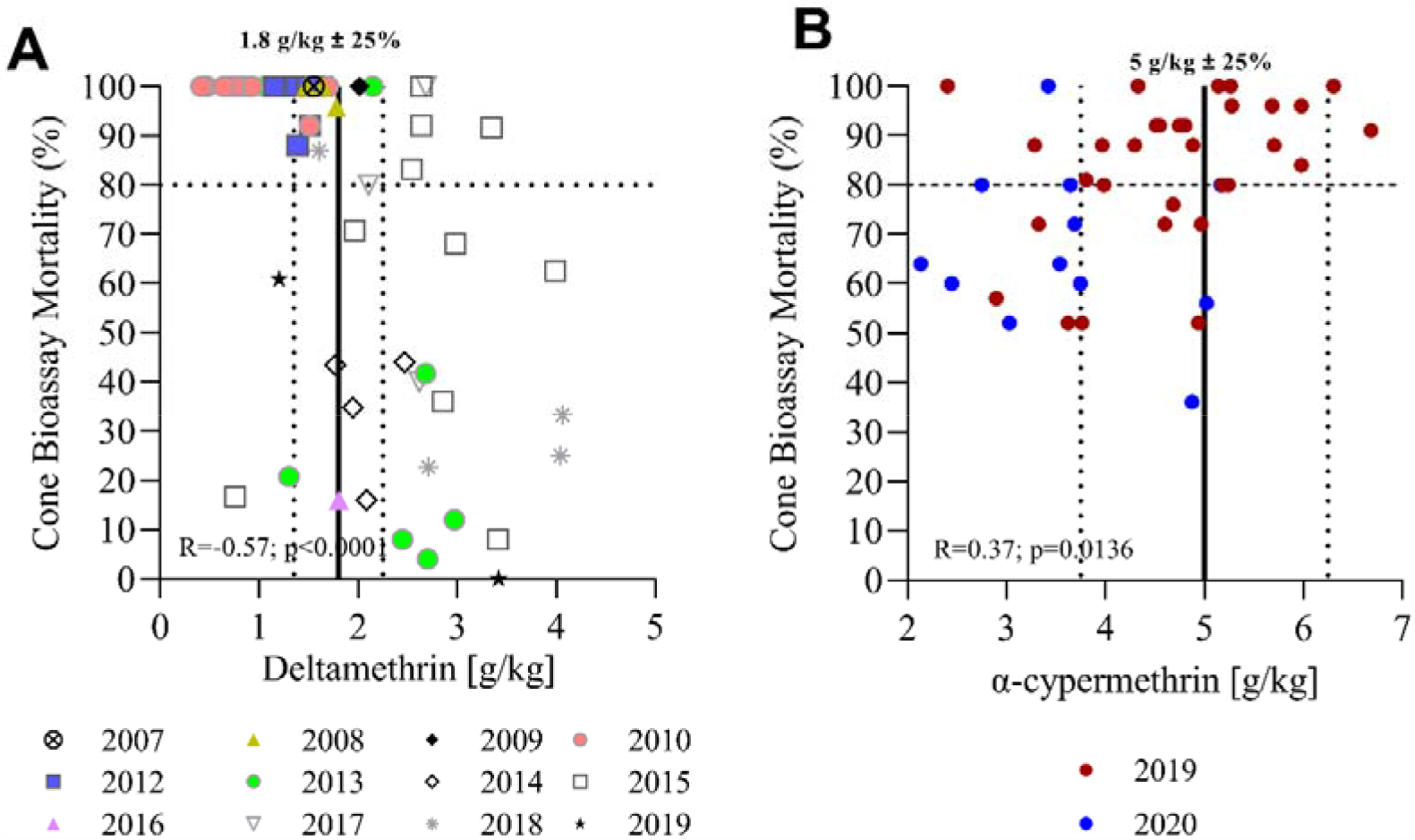
Correlation of total insecticide content and bioefficacy. **Panel A** shows the correlation of total AI content as measured by LC-MS with mortality rates observed in standard cone bioassays for PermaNet® 2.0. **Panel B** shows the same data for SafeNet®. Vertical lines indicate the target AI concentrations as per product label (average +/-25%). The horizontal line indicates the 80% threshold mortality for standard WHO cone bioassays.

## Discussion

The present study was aimed at estimating the accuracy of XRF, an in-field tool to quantify not only deltamethrin but also alpha-cypermethrin in LLIN samples. For this, LLIN samples were obtained from consignments of LLINs for mass distribution in Papua New Guinea (PNG). A robust method to quantify insecticide content may be beneficial for researchers and programs, e.g., for use in LLIN durability studies or for in-country quality assurance spot checks. These results illustrate that XRF and LC-MS data are highly correlated indicating that XRF could serve this purpose. However, the Bland and Altman analyses show that the agreement between methods in this study was subject to substantial uncertainty. The major source for this uncertainty is that samples (while derived from the same LLINs), were not exactly the same, i.e., the XRF measurements were done on a different (adjacent) part of the same net as compared to the LCMS measurements. It is known that individual LLINs can exhibit substantial spatial variation in insecticide content as a result of the manufacturing process [27]. As such, the observed uncertainty is not surprising. Despite this, these results showed that even single measurements on different samples from the same LLINs using the two different methods are very well correlated on average. Further studies averaging multiple XRF and LCMS measurements across single LLINs are needed to assess the accuracy of XRF to predict if a product conforms to label specifications.

The CIPAC methods use HPLC in quantifying the pyrethroids and utilize a variety of extraction conditions depending on the insecticide/fibre combination (CIPAC, 2009 #61;CIPAC, 2012 #60). In contrast, the method presented here employs a simple extraction, using methanol-only as solvent and without heat to prepare samples for LC-MS. The data presented here provide evidence that the method is robust. Both, solvent volume and extraction time did not influence results significantly, and extraction was shown to be complete (>95%) after one extraction step. This method is unlikely to be directly transferrable to incorporated nets but may be suitable for other coated net products. Methanol causes the chemical isomerization of deltamethrin [28]. As such, this study protocol (limited by the availability of equipment) did not allow for an analysis of the deltamethrin isomers present in the samples. Further studies are required to confirm the isomer ratio of deltamethrin in PermaNet® 2.0 LLINs manufacture prior-or in 2012 and beyond 2012.

The second objective of this study was to correlate bioefficacy results as measured in standardized WHO cone bioassays with total AI content measurements. This was done to explore if total AI content is a robust indicator of bioefficacy in new and unused LLINs. With PermaNet® 2.0, the authors observed an inverse correlation between bioefficacy and total AI content which is unexpected and counterintuitive, indicating that total AI concentration measurements do not always predict bioefficacy well. The reason for decreased bioefficacy of most of the recent PermaNet® 2.0 (2013 - 2019) can be explained by a change in the coating formulation which causes restricted bioavailability on the net surface leading to reduced mosquito mortality as observed in WHO cone bioassays [5].

### Conclusions

This study showed that XRF is a promising in-field method not only for deltamethrin but also for alpha-cypermethrin. Further studies should investigate XRF utility for measuring other insecticides used in the public health space (e.g., for indoor residual spraying). This study provided evidence that total AI concentration is not a robust indicator of bioefficacy in new and unused LLIN products, and bioefficacy tests should be included in predelivery inspections.

For PermaNet® 2.0, the nets with high AI content performed poorly in standardized cone bioassays.

## Data Availability

All data produced in the present study are available upon reasonable request to the authors

## List of abbreviations

AI: Active ingredient
CIPAC: Collaborative International Pesticides Analytical Council
HPLC: High-pressure liquid chromatography
IMR: Institute of Medical Research
LC-MS: Liquid chromatography mass spectrometry
LLINs: Long-lasting insecticidal nets
PNG: Papua New Guinea
RAM: Rotarians Against Malaria
WHO: World Health Organization
XRF: X-ray fluorescence

## Declarations

### Ethics approval and consent to participate

Not applicable.

### Consent for publication

Not applicable.

### Availability of data and materials

The data set for this study is available on request from the corresponding author.

### Authors’ contributions

Conceived Study: SK; Conducted XRF analysis and Bioassays: NB, EA; Conducted LC-MS: DW, MK; Wrote first draft: SK, MK; Data Analysis: SK, MK; Reviewed Manuscript: All Authors.

## Acknowledgement

The authors thank the PNG National Malaria Control Program (Leo Makita) and Rotarians Against Malaria PNG (Tim Freeman) for making available the LLIN samples for testing. We would also like to thank William Pomat, Moses Laman and the staff of the PNGIMR Vector-borne Diseases Unit for operational support.

